# Determinants of insecticide treated bed nets use among pregnant women in the Kintampo north municipality, Bono East region, Ghana

**DOI:** 10.1101/2025.05.13.25327488

**Authors:** Issah Sumaila, Shaibu Issifu, Mubarick Nungbaso Asumah, Kwaku Nimo-Boakye, Debora Appietuah Awuah, Helen Agodzo, Gavin Apio, Anthony Twum

## Abstract

Malaria is an avoidable ailment of public health importance with increased morbidity and mortality. Insecticide-treated nets (ITNs) are proven defence intended to limit contacts with mosquitoes thereby preventing malaria morbidities and mortalities. The aim of this study is to assess the determinants of regular ITN use among pregnant women in the Kintampo North Municipality.

A facility-based descriptive cross-sectional study was employed in the Kintampo North Municipality. Pregnant women who sought antenatal care services were recruited. Data was collected with the aid of structured questionnaire with respondents recruited using random sampling method. A P-value of < 0.05 was considered as statistically significant.

Almost all (98.1%) respondents were aware of ITNs and majority (85.4%) indicated that ITNs are meant for protection against mosquito bites. Half (53.5%) of respondents heard about ITNs from hospitals/clinics. Majority (73.9%) of respondents identified health facilities as the venue to access ITNs. ITN usage, mosquito repellent, keeping surroundings tidy were ways reported to prevent malaria. Barriers to regular usage of ITN were skin rashes and cumbersome to use. Being married (aOR:3.9, 95%CI:1.64-9.28), hanging bed nets (aOR:17.8, 95% CI: 7.07-44.8) and willing to buy ITNs (aOR: 2.06, 95%CI: 1.05-4.04) were the key determinants of regular ITNs use.

There is high awareness and knowledge about the strategies to avoid malaria using ITNs. There is the need for continuous sensitization. There should be community engagements by the Ministry of Health and other stakeholders to help eschew all myths and misconceptions about the use of ITN.

## Introduction

Malaria is a preventable illness of public health importance with high annual morbidity and mortality rate. Malaria is a devastating parasitic infectious disease, killing over one million people annually [1]. All pregnant women are at risk of developing malaria at some point in their peri-conception periods [2]. Despite years of global efforts to combat malaria, the disease continues to exact a devastating toll, with mortality rates remaining stubbornly high at 435,000 deaths per year between 2015 and 2017 [3]. Organizations such as Partnership to End Malaria, Roll Back Malaria (RBM), African Union Commission and other partner organizations join the WHO in promoting “Zero Malaria Starts with me,” a primary campaign that targets at rating malaria high on the political agenda, mobilizing additional resources, and empowering communities to take ownership of malaria prevention and care in malaria endemic countries [4]. Malaria in general is a serious global public health issue, affecting over 3.3 billion people [5]. Over 50% of the pregnant women who report at the antenatal clinic are malaria positive [6,7]. Globally, 585,000 women die in developing countries from malaria in pregnancy related complications [8]. Indoor Residual Spraying (IRS) and Insecticide Treated bed Nets (ITNs) have been recommended as the most effective preventive measures that can reduce contact between mosquitoes and humans through their insecticidal effect and providing barrier respectively. A report by WHO showed that ITN coverage increased slightly and protected about half of all the people at risk of malaria in Africa from 2015 to 2017 compared to 29% in 2010 [9]. Despite these interventions, approximately 12 million dollars is spent annually in diagnosing and treating malaria with an approximate economic loss of 1.3% in highly endemic countries [10].

In sub-Saharan Africa (SSA), malaria in pregnancy is a major contributor to adverse pregnancy outcomes including intrauterine-growth retardations, low-birth-weights, stillbirths, spontaneous abortions, preterm deliveries, severe anaemia and maternal deaths [6]. Malaria’s impact on pregnancy miscarriages in sub-Saharan Africa is poorly understood, including the mechanisms of transmission, intensity of infection, and development of protective immunity among pregnant women, which affects the risk of complications from antenatal malaria infections [6,11]. This is because it has not been well documented. Malaria parasites sequester and replicate in the placenta making pregnant women more susceptible and three times more likely to be infected with malaria than non-pregnant women being infested with malaria parasites from the same area [6,11,12]. Sub-Sahara Africa has the greatest burden of malaria during pregnancy with a stable malaria transmission [13]. A characteristic of malaria infection among pregnant women in stable transmission areas is that it is often asymptomatic due to the pre-existing immunity that has been acquired through frequent exposure to *P. falciparum* malaria infections since childhood [14]. The absence of clinical symptoms such as fever makes it difficult to diagnose the disease and thus often remain untreated until it causes complications [15]. A study that was conducted in Nigeria to investigate malaria parasitaemia among pregnant women reported a 59.9% prevalence of malaria infection. The highest prevalence occurred in the first trimester (84.1%). Among the positive cases, mild parasitaemia, moderate parasitaemia and severe parasitaemia were respectively 47.2%, 37.4% and 15.3% of the total cases [16]. Like the other Sub-Saharan African countries, malaria in Ghana is endemic and the entire population is at risk of malaria infections. The incidence of malaria still accounts for 40.0% of all outpatient department attendance, with the most vulnerable groups being pregnant women and children under five years of age [17]. Ghana’s malaria transmission is heterogeneous and differs along varying ecological zones. Parasite prevalence is seasonal, peaking in the wet season (June–October) in the northern savannah area. However, in both forests and coastal ecological zones, malaria parasite prevalence peaks twice annually [18].

In Ghana, the death of a pregnant woman is considered a tragic event but most of the maternal deaths are caused by malaria infections among pregnant women which recently accounts for about 28.1% of Out-Patience Department (OPD) attendance, 13.7% of hospital admissions and 9.0% of maternal deaths[19, 20]. According to the 2018 Annual Health Report of the Kintampo North Municipality, 178 (23.7%) of the 750 pregnant women who visited the Kintampo Municipal Hospital were infected with malaria [21]. All pregnant women attending antenatal clinic are given free ITNs as a strategy to reduce malaria yet, malaria increased by 9.1% [22]. This implies that, other factors expose pregnant women in the Kintampo Municipality to malaria infection. This study, therefore, aimed at identifying the determinants for the regular use of ITNs among pregnant women attending the Kintampo Municipal Hospital.

## Methods

### Study area

The study was conducted at the Kintampo Municipal Hospital (KMH). This facility is the only government Hospital in the Municipality. This study setting was chosen because, KMH serves as a referral facility for all 23 facilities within the Kintampo North Municipality in the Bono East and even neighbouring districts.

### Study design

This was a facility-based descriptive cross-sectional study with emphasis on the quantitative research approach. Respondents were selected within a period of 8 weeks (4^th^ May, 2024 to 2^nd^ July, 2024) at Kintampo Municipal Hospital.

### Study population

The study population were pregnant women who attended Kintampo Municipal Hospital’s ANC during the time of data collection.

### Inclusion and exclusion criteria

All pregnant women in all trimesters who consented to participate in the study were included in the study. Pregnant women who met the inclusion criteria but did not consent to participate in the study were excluded.

### Sample size determination, sampling method and procedure

The sample size was calculated using Yamane’s formula [23]. The average monthly attendance at the KMH antenatal was 1200 pregnant women. A 5% margin of error with 95% confidence level was used to calculate the sample size. The sample size was thus estimated as 300. A 5% of the calculated sample size was added to make up for the non-response rate. The final the sample size was estimated as 315.

A total of 315 pregnant women were subsequently recruited in the study, from the Kintampo Municipal Hospital antenatal clinic. The ANC operates on Mondays to Fridays every week. Pregnant women attending the antenatal clinic at Kintampo Municipal Hospital during the period of this study constituted the study population. The study employed simple random sampling technique. The approximate 1200 pregnant women were estimated to have attended the hospital in previous months, representing the number of the entire study population. Patients who presented at the ANC on each day of data collection were randomly selected using the balloting method. In this method, pieces of paper were used on which either ‘Yes’ and ‘No’ were written, folded and put in a bowl. Patients who picked ‘Yes’ were included in the study after they had provided informed consent whereas those who picked ‘No’ were automatically excluded. This procedure was repeated until the desired sample size was achieved over a period of about 12 weeks.

### Data collection Tool and Procedure

Structured questionnaire was designed based on the study objectives to elicit information from the respondents. The instrument translated into local languages for those without formal education. The questionnaire was made of seven (7) sections: Section A constitutes demographic and socio-economic characteristics of the respondents. Section B constituted their knowledge level on how to prevent malaria during pregnancy, Section C elicited information on respondents’ level of knowledge on in ITNs. Section D discussed ownership of ITNs by respondents, Section E constituted their willingness to purchase ITNs, Section F looked at how to improve coverage and Section G discussed whether respondents encountered problems while using ITNs.

### Data Analysis and Presentation

Data was entered, cleaned and managed using Microsoft Excel version 16. The data was subsequently exported to Stata version 15 and analysed. Gender, religion, marital status, and educational level were coded as categorical variables. Age and household size were collected as continuous variables and categorized during analysis. Descriptive statistics were used to present the proportions on background characteristics of the respondents at the univariate level. Regular utilisation of ITNs for this study was defined as sleeping under the treated ITN the night preceding the study [24]. This was followed by a bivariable analysis using Pearson’s Chi-square test of independence to test for association between the dependent and independent variables. Significance level was pegged at p<0.05.

Multivariable logistic regression was fitted after the bivariable analysis. The multivariable logistic regression was done at two levels. The first was to establish the crude level of association between the independent variables and the dependent variable for categorical variables. In this strategy, all independent variables were added to the first model to establish the crude relationship with the dependent variable at various levels of measurement. Afterwards, variables that were significant at the bivariable analysis level were entered in a multivariable logistic regression model to adjust for the confounding effects of those variables. Tables and graphs were used to present the results.

### Ethical Issues

Ethical clearance approval was granted by Committee on human research, publication and ethics (CHRPE) of Kwame Nkrumah University of Science and Technology, Kumasi for the study with clearance certificate number **CHRPE/AP/324/24**.

## Results

### Socio-Demographic and Socioeconomic Characteristics of Respondents

In all, 315 respondents who took part in the study where, higher proportion (32.1%), were within 20 - 24-year age group with a mean age and standard deviation of 26.1± 5.5 years. Also, the study revealed that 43.5% of the respondents were in their second trimester. Christianity is the dominating religion within the study area (61.6%). The study indicated that 73.3% of the respondents were married while only 1.0% were widowed. More than half (53.4%) of the respondents had a household size of four or more. The study revealed that those who attained JHS certificate were slightly higher than the remaining levels with 26.8% whilst primary education was the least representing 15.3%. Moreover, place of residence was almost equally distributed, 54.6% reside in the urban communities while the 45.4% leave in rural areas. On occupation, the findings indicated that 36.8% were traders whereas only 4.8% were not among the occupation types listed. A greater proportion of the study participants, 77.9% earn a monthly income of <500 GHS whereas only 9.0% receive amount >GHS 1000 monthly.

### Knowledge of respondents on the use of ITNs

The study revealed that almost all the respondents 98.1% have heard about ITNs before. Of the 98.1% of the respondents who ever heard of ITNs, majority 85.4% indicated ITNs are used for protection against mosquito bites. Among those who ever heard about ITNs, slightly more than half 53.5% indicated hospital/clinic as their main sources of information. The study further revealed that, majority 73.9% of the respondents identified health facilities as where to access ITNs whereas 9.5% said ITNs can be accessed from community volunteers. Also, a greater proportion of the study population 62.1% indicated they are not aware that ITNs can be retreated whilst 37.9% confirmed that it could be retreated.

### Barriers to ITNs use among pregnant women the Kintampo Municipality

Table 3 below presents some of the factors that impede the use of ITNs among pregnant mothers within the Kintampo Municipality. When respondents were asked whether their ITN is currently hanged or not, 80.4% affirmed it is being hanged. Among the reasons for not hanging ITNs, majority (59.0%) were Allergic to use ITN. On the question of whether or not respondents slept under ITN the previous night, 28.9% responded negatively as did not sleep under ITN. The major reason for not sleeping under ITN the night before was feeling hot inside ITNs (55.5%). The results further indicated that 41.4% of the respondents had no willingness to buy bed nets. Among the reasons for the unwillingness, majority 40.8% indicated they are financially constrained whereas a few (7.7%) said bed nets look like burial shroud. Also, when respondents were asked whether or not they have problems with the use of the ITN, slightly less than half of the respondents responded affirmatively. The main barrier for not using ITN was, it produces skin rashes (49.7%) whilst a few responses gathered as it is cumbersome to use (11.3%).

**Table 1:**
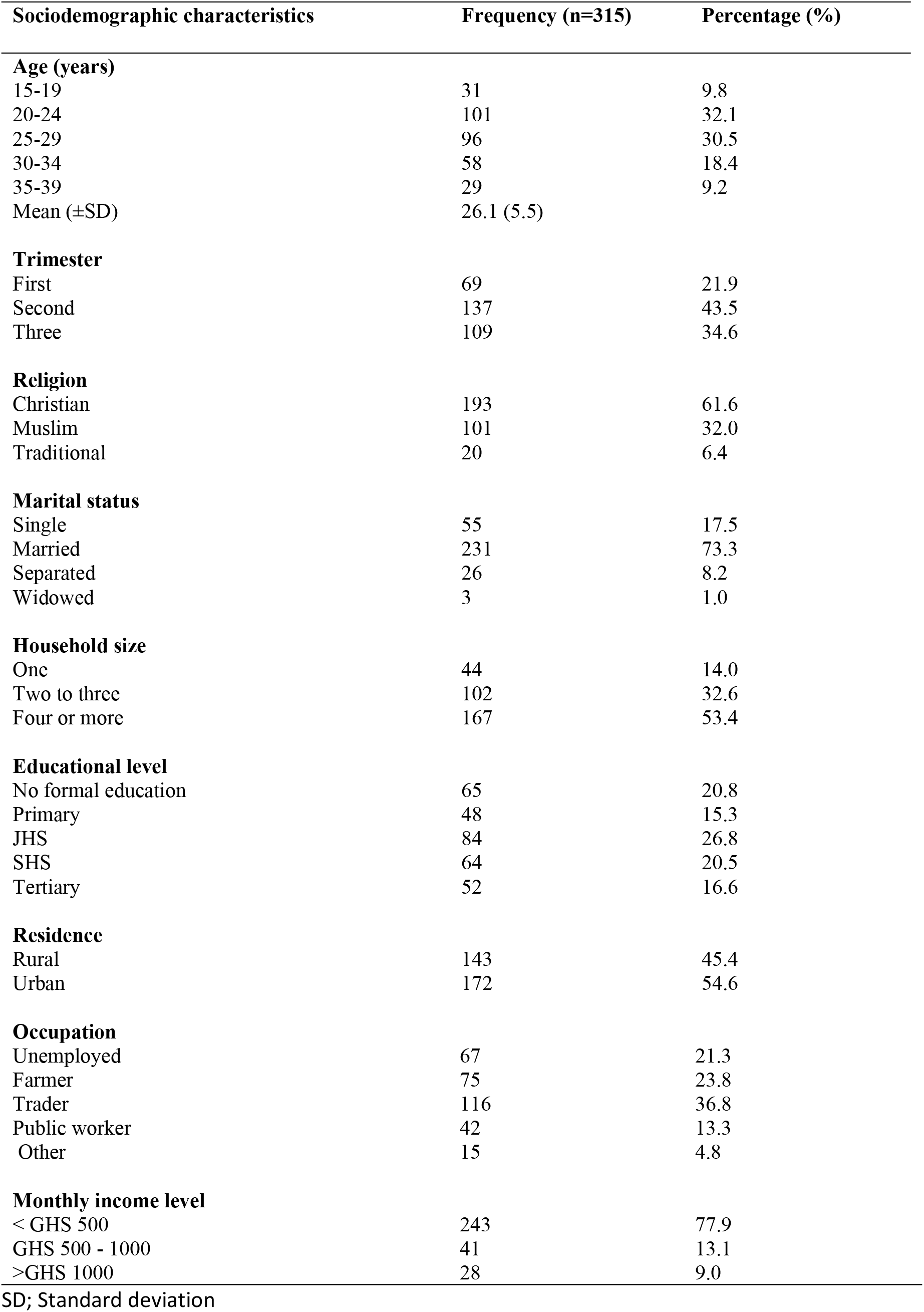
Sociodemographic characteristics of respondents.

**Table 2:** Respondents knowledge on the use of ITNs.

| **Knowledge on malaria and ITN use** | **Frequency (N=315)** | **Percentage (%)** |
| --- | --- | --- |
| **Heard about ITNs** |  |  |
| No | 6 | 1.9 |
| Yes | 309 | 98.1 |
| **Uses of ITNs** |  |  |
| Room decoration | 26 | 8.4 |
| Protection against mosquito bites | 264 | 85.4 |
| Affording good sleep | 14 | 4.5 |
| Fishing | 5 | 1.6 |
| **Sources of information about ITNs** |  |  |
| Television | 45 | 14.3 |
| FM station | 27 | 8.6 |
| District Health Education | 53 | 16.9 |
| Hospital/clinic | 168 | 53.5 |
| Relatives/Peers | 15 | 4.8 |
| Other | 6 | 1.9 |
| **Where to access ITNs** |  |  |
| Community volunteers | 30 | 9.5 |
| Health facilities | 232 | 73.9 |
| Pharmacy shops | 52 | 16.6 |
| **Awareness that ITNs can be re-treated** |  |  |
| No | 195 | 62.1 |
| Yes | 119 | 37.9 |

**Table 3:** Barriers to ITNs use among pregnant women in the Kintampo Municipality.

| Variables | Frequency (N = 315) | Percentage (%) |
| --- | --- | --- |
| **Your ITN is currently hanged** |  |  |
| No | 61 | 19.6 |
| Yes | 251 | 80.4 |
| **Reasons for not hanging ITN** |  |  |
| Not part of topmost priority | 15 | 24.6 |
| Allergic to use ITN | 36 | 59.0 |
| Looks like burial shroud | 10 | 16.4 |
| **Slept under ITN the previous night** |  |  |
| No | 90 | 28.9 |
| Yes | 222 | 71.1 |
| **Reasons for non-use of ITNs** |  |  |
| Feels hot inside | 50 | 55.5 |
| Allergic to chemicals in ITN | 4 | 4.6 |
| Looks like burial shroud | 13 | 14.9 |
| Other | 20 | 23.0 |
| **Willingness to buy bed net** |  |  |
| No | 130 | 41.4 |
| Yes | 184 | 58.6 |
| **Reasons for unwillingness to buy ITNs** |  |  |
| Financial constraint | 53 | 40.8 |
| Not part of topmost priority | 46 | 35.4 |
| Allergic to use of ITN | 21 | 16.1 |
| Looks like burial shroud | 10 | 7.7 |
| **Having problems with the use of the ITN** |  |  |
| No | 162 | 51.8 |
| Yes | 151 | 48.2 |
| **Barriers to bed net use** |  |  |
| Skin rashes | 75 | 49.7 |
| Generates heat | 59 | 39.0 |
| Cumbersome to use | 17 | 11.3 |

### Motivators to ITNs use among pregnant women the Kintampo Municipality

Majority (71.1%) of the respondents indicated they slept under ITN the previous night. Most of them (82.6%) slept under the ITNs to prevent mosquito bites. Also, 63.0% of the respondent uses

ITNs all year round of which the majority (85.9%) reason was to prevent malaria. Again, majority (84.3%) of the respondents stated that the fundamental benefit to ITN usage is to prevent malaria. Among the ways to increase ownership of ITNs, a significant number (38.5%) of the respondents suggested that the nets should be made free, whilst community education on ITN use and regular ANC attendance were also indicated by 26.1% of the pregnant women as other way of increasing ownership of ITNs. The study further revealed free distribution of ITNs (63.9%) as a major motivator to the use of ITN among pregnant mothers. Further details are presented in table 4.

**Table 4:** Motivators to ITNs use among pregnant women in the Kintampo Municipality.

| **Variables** | **Frequency (N=315)** | **Percentage (%)** |
| --- | --- | --- |
| **Slept under ITN the previous night** |  |  |
| No | 90 | 28.9 |
| Yes | 222 | 71.1 |
| **Reasons for use of bed net** |  |  |
| Feels cold inside | 6 | 2.7 |
| For privacy | 15 | 6.7 |
| To prevent mosquito bites | 185 | 82.6 |
| Habitual | 8 | 3.6 |
| Advised by a midwife to use ITN | 9 | 4.0 |
| Other | 1 | 0.4 |
| **Period of using ITNs** |  |  |
| All year round | 196 | 63.0 |
| During rainy season | 79 | 25.4 |
| During dry season | 9 | 2.9 |
| Other | 27 | 8.7 |
| **Reasons for using ITN during season** |  |  |
| To prevent malaria | 267 | 85.9 |
| For warmth | 19 | 6.1 |
| As a partition in the room | 1 | 0.3 |
| To provide privacy | 8 | 2.7 |
| Other | 16 | 5.1 |
| **Benefits of using ITN** |  |  |
| To prevent malaria | 263 | 84.3 |
| To sleep soundly | 20 | 6.4 |
| To provide warmth | 7 | 2.2 |
| To prevent insects bite | 18 | 5.8 |
| Other | 4 | 1.3 |
| **Ways to increase ownership of ITNs** |  |  |
| Reduce price | 25 | 8.0 |
| Community education on ITNs use | 82 | 26.1 |
| Regular ANC attendance | 82 | 26.1 |
| Regular CWC attendance | 2 | 0.6 |
| Nets should be free | 121 | 38.5 |
| Other | 2 | 0.6 |
| **Ways to improve ITNs use** |  |  |
| Free distribution of ITNs | 198 | 63.9 |
| Intensive education on ITNs | 89 | 28.7 |
| Modification of ITNs | 18 | 5.8 |
| Other | 5 | 1.6 |

### Determinants of regular ITNs use of respondents and its association with sleeping under the ITNs

The odds of determinant of regular ITNs use for married respondents was 3.9 times compared to that of those who were single (p-value=0.002, CI=1.64 – 9.28). Again, the odds of determinant of regular ITN use among respondents who hanged their ITNs is 17.8 times compared to those who did not (p < 0.001, CI=7.07-44.76). Findings from the study further revealed that respondents who were willing to purchase ITNs were 2.1 times more likely to be regular users compared to those were not willing (p=0.036, CI=1.05 – 4.04). Details are as shown in the table 5 below.

**Table 5:** Determinants of regular ITNs use of respondents and its association with sleeping under the ITNs.

| Variables | Unadjusted |  |  | Adjusted |  |  |
| --- | --- | --- | --- | --- | --- | --- |
|  | uOR | 95% CI | P-value | aOR | 95% CI | P-value |
| **Age (years)** |  |  |  |  |  |  |
| 15-19 | Ref |  |  | Ref |  |  |
| 20-24 | 1.32 | 0.57 - 3.05 | 0.513 | 1.11 | 0.36 - 3.36 | 0.858 |
| 25-29 | 1.89 | 0.80 - 4.47 | 0.144 | 1.26 | 0.40 - 3.92 | 0.697 |
| 30-34 | 2.70 | 1.02 - 7.16 | **0.046\*** | 1.72 | 0.45 - 6.55 | 0.427 |
| 35-39 | 0.98 | 0.80 - 4.47 | 0.964 | 0.50 | 0.12 - 2.07 | 0.335 |
| **Religion** |  |  |  |  |  |  |
| Christian | Ref | - | - | Ref | - | - |
| Muslim | 0.95 | 0.55 - 1.631 | 0.852 | 0.76 | 0.36 - 1.62 | 0.479 |
| Traditional | 0.30 | 0.12 - 0.76 | **0.011** | 1.08 | 0.31 - 3.84 | 0.901 |
| **Marital status** |  |  |  |  |  |  |
| Single | Ref | - | - | Ref | - | - |
| Married | 3.08 | 1.67 - 5.71 | **<0.001** | 3.90 | 1.64 - 9.28 | **0.002\*** |
| Separated | 1.75 | 0.67 - 4.62 | 0.255 | 3.09 | 0.80 - 11.95 | 0.102 |
| Widowed | 0.46 | 0.04 - 5.43 | 0.541 | 0.87 | 0.03 - 24.83 | 0.934 |
| **Uses of ITNs** |  |  |  |  |  |  |
| Room decoration | Ref | - | - | Ref | - | - |
| Protection against mosquito bites | 3.09 | 1.36 - 7.02 | **0.007** | 2.47 | 0.85 - 7.20 | 0.097 |
| Affording good sleep | 1.8 | 0.47 - 6.85 | 0.389 | 1.80 | 0.27 - 11.97 | 0.543 |
| **Sources of information about ITNs** |  |  |  |  |  |  |
| Television | Ref | - | - | Ref | - | - |
| FM station | 3.85 | 1.24 - 11.97 | **0.02** | 5.54 | 0.99 - 30.93 | 0.051 |
| District Health Education | 2.16 | 0.93 - 4.99 | 0.072 | 0.94 | 0.31 - 2.88 | 0.918 |
| Hospital/clinic | 3.18 | 1.59 - 6.36 | **0.001** | 2.38 | 0.93 - 6.05 | 0.069 |
| Relatives/Peers | 0.58 | 0.178 - 1.91 | 0.374 | 1.41 | 0.22 - 9.07 | 0.716 |
| Other | 0.44 | 0.07 - 2.64 | 0.367 | 0.78 | 0.07 - 8.57 | 0.841 |
| **Your ITN is currently hanged** |  |  |  |  |  |  |
| No | Ref | - | - | Ref | - | - |
| Yes | 20.92 | 10.24 - 42.73 | **<0.001** | 17.79 | 7.07 - 44.76 | **<0.001\*** |
| **Willingness to buy bed net** |  |  |  |  |  |  |
| No | Ref | - | - | Ref | - | - |
| Yes | 3.362 | 2.02 - 5.60 | **<0.001** | 2.06 | 1.05 - 4.04 | **0.036\*** |

**\*Significant (p<0.05); uOR = unadjusted odds ratio; aOR = adjusted odds ratio; Ref =** **reference**

## Discussion

This study aimed to identify the determinants for the regular use of ITNs among pregnant women attending Kintampo Municipal Hospital. The findings of the current study showed that almost all pregnant women have heard about ITNs (98.1%). This is consistent with other studies conducted in Ghana. For instance, a qualitative study conducted by Manu et al., [25] in the middle belt of Ghana showed that all pregnant women were said to have heard about ITNs. In Wa (Upper West Region, Ghana), 95.9% of pregnant women have heard of ITNs [26], in the Hohoe municipality (Volta Region, Ghana), 98.7% of pregnant women were aware of ITNs, in the Kassena-Nankana East Municipality, (Upper East Region of Ghana), 99.7% of pregnant had heard about ITNs [27]. In comparison with other studies in different jurisdictions, the awareness of ITNs among pregnant women in the current study is higher than 76.1% reported in Northern Uganda [28] and 93.2% in Nigeria [29]. In Ghana, malaria is among the top ten (10) disease conditions [17]. The consequences of malaria on pregnant women and children under five are dire [27]. As such, there are deliberate interventions to disseminate accurate information on bed nets to influence the use of ITNs. This therefore could account for the higher awareness reported in these studies.

More than half (53.0%) and about 25.0% of the respondents got information on ITNs through the hospital/clinics and media (TV and Radio) respectively. The study is consistent with that of Amara [30] whose study showed that majority of study participants have heard of ITNs from the health care workers. In Nigeria, most (45.0%) pregnant women had information about ITNs at antenatal care (ANC) [31]. This is because, at the ANC units, pregnant women are educated on diseases common in pregnancy, including malaria and their prevention. In our setting, the simplest way to prevent malaria is to use ITNs [9]. That is why as much as possible every pregnant woman is given a treated bed net as soon as pregnancy is confirmed. This, therefore, explains the similarities. Also, about a quarter of the respondents had heard of ITNs via the media. Contrary, Nungbaso et al., [32] have shown that most respondents have heard of ITNs via the media. The difference in demographic characteristics of the study participants could influence the disparities in the results. Often times, there is very limited time for education of conditions at the ANC as a result of workload. In the Kintampo Municipality, the public health unit visits the radio stations twice every week to educate the population on disease conditions and their prevention. With the radio talk shows, these women are comfortable in their homes and listen attentively. Also, they can call into the program to seek clarity. Information dissemination on ITNs is very essential to its usage. That explains why a quarter of the respondents indicated that they have heard of ITNs. To meet a higher audience, there is the need to vary the sources of information on ITNs.

About eight in every ten respondents knew that ITNs were used to prevent malaria or mosquito bites. This is similar to a study conducted in Northern Ghana where a higher proportion of respondents knew the actual use of bed nets [32]. This is also essential in the utilization of ITNs because if people do not believe in the purpose and effectiveness of the ITNs, they are likely not to use them. The ability of the study participants to acknowledge the right use of ITNs means that the education on malaria prevention is getting some desired results.

Only 37.9% knew that ITNs could be retreated. Similarly, about 70.0% of residents in Kampala, Uganda were not aware that ITNs could be retreated [33]. Retreatment of bed nets is done by dipping the nets in a water solution containing insecticides and then drying them in the shade [34– 36]. However, the frequent treatment of ITNs was seen as a barrier to the utilization of ITNs.

Currently in Ghana retreatment of bed nets is substituted with the frequent free distribution of bed nets.

The utilization of ITNs was found to be 71.1% in this study. This is higher than 69.3% reported in Wa [26], 66.4% of pregnant women in Hohoe municipality reported ITNs usage [37]. Also, in Ethiopia, 70.8% of pregnant women use ITNs [38]. Though the utilization of ITNs as reported in this study is considered high, it is lower than the 74.2% average ITNs utilization reported in a multilevel study across 21 Sub-Sahara African countries [39]. The slight difference could be attributed to the geographical location and demographic characteristics of the study participants. Contrary to the above, the utilization of ITNs in this study is higher than the 35.0% utilization rate of ITNs among pregnant women in Uganda [28] and 19.2% reported in Nigeria [29]. The difference could be explained as follows; in Ghana, the Ministry of Health (MoH) through Ghana Health Services (GHS) with the support of agencies including but not limited to UNICEF and WHO has intensified the free distribution of ITNs. Also, the Kintampo Health Research Centre (KHRC) has carried out a lot of interventions on malaria prevention especially among pregnant women and children under five. These interventions in Ghana could account for the difference. It is worthy to note that ownership of ITNs does not translate to usage[32]. As such, it is necessary to go beyond just the free distribution of bed nets. There should be multiple strategies aimed at increasing ITNs usage.

In relation to the above, the study observed that aside the usage of ITNs, some respondents also use other preventive methods such as the use of mosquito repellents, mosquito sprays, weeding, and cleaning the environment. Thus, the remaining 28.9% of respondents who did not use ITNs could still be using other preventive measures to prevent malaria in their households.

In the current study, the majority of the study participants hung their ITNs. The study further revealed that those who hang their bed nets were likely to use them (AOR=17.8, p<0.001). Hanging the bed nets is a good step in using ITNs. This is because in this study some people cited being lazy to hang their ITNs as a reason for not using them. This is related to a study in South-Eastern Nigeria where 69.1% hung bed nets over the bed and slept under it after tucking it underneath the mattress as well as avoiding contact with the body[31]. Though assessing the correct use of ITNs is essential to measure the progress of the sensitization program, the current study could not explore that aspect. [31] further argued that most individuals are now aware of their role in the use of ITNs to achieve the full benefits of ITNs. This is a serious aspect of ITNs utilization that requires more research.

Out of those who did not hang their ITNs, 59.0% believed they were allergic to the chemical in ITNs with 49.7% reporting skin rashes upon using ITNs. This is related to several studies [7,15– 17] where adverse reactions, skin irritations are commonly cited as the reason for not using ITNs.

Among those who were not using ITNs, 55.5% cited being hot in ITNs as their reason. This is similar to other studies conducted elsewhere. For example, in Nigeria, 74.4% of pregnant did not use ITNs citing excessive heat and discomfort as their reasons [6]. A qualitative study in Ghana also showed that the common reason mentioned by respondents for not using ITNs was heat [15]. In Myanmar, feeling hot is cited as the reason for not using ITNs [18,19]. This necessitates behaviour change communication (BCC) tactics to include effective health messaging and advertisement that explain ITNs benefits, common myths and misconceptions about it, the adverse reactions, and the fact that these side effects (i.e., skin rashes, etc.) are only temporary.

The study showed that married pregnant women were more likely to use ITNs as compared to their counterparts who were single. It could be that the married women get some support from their husbands. A husband who cares so much about her pregnant wife and follows his wife to the ANC would understand the need to sleep under ITNs. The husband who is informed about the importance of ITNs would serve as security in ensuring that the health of the child and the mother are safeguarded by sleeping in the ITNs. In this light, we call for more sensitization on the need for husbands to accompany their wives for the ANC visits. This will enable the men to check whether their wives are adhering to the teachings at ANC.

Also, willingness to buy ITNs was a key determinant to ITN usage. This is related to other studies where increase wealth increases the likelihood of using ITNs [27,40,41]. Pregnant women with average or more wealth quintiles can feed their families and maybe be able to save some money to buy other basic needs. The poorest person may be struggling to feed the family as such may not prioritize sleeping in ITNs to being hungry. To increase utilization of ITNs, there is the need to devise means to ensure all persons irrespective of their locations, place of work, etc. benefit from the government free distribution of ITNs.

## Limitations of the Study

The study was cross sectional study as such no cause-effect relationship could be established. The study participants included only expectant mothers who were attending ANC, as such generalization of these findings ought to be done with caution. The sample size was not large for some factors that were found to be significant hence they had wider confidence interval.

## Conclusions

The study accounts for higher awareness of ITNs among pregnant women in the Kintampo North Municipality, nevertheless, this could not adequately decipher into the utilization of the ITNs with varied reasons given by the pregnant women. This calls for prompt diverse strategies to be rolled out to encourage the utilization of bed nets beyond their free distributions. This could be championed by periodic sensitizations and education by the Ministry of Health (MoH) and its agencies and private bodies including UNICEF to help eschew all forms of myths and misconceptions about the use of ITNs. To increase awareness of ITNs, there is the need to vary the information sources so as to reach out with a higher audience with the information on ITNs and its usage. Significant efforts and attentions should be tailored towards the eradication of the recognized determinants that obstruct the utilization of ITNs in the municipality and the region as a whole.

## Data Availability

The data has bee uploaded as part of this submission

## Acknowledgements

We acknowledge the support and encouragement of the management of Kintampo Municipal Hospital throughout the study.

## Supporting information

S1_file.docx. **This the Strengthening the Reporting of Observational Studies in Epidemiology (STROBE) checklist**

